# A qualitative study exploring the diagnostic and treatment journeys of children and young people with gastroduodenal disorders of gut-brain interaction, their families, and the clinicians who care for them

**DOI:** 10.1101/2024.10.15.24315420

**Authors:** Gayl Humphrey, Mikaela Law, Celia Keane, Christopher N Andrews, Armen Gharibans, Greg O’Grady

## Abstract

**Background:** Gastroduodenal disorders of gut-brain interaction (DGBI) are prevalent in the paediatric population. Diagnostic pathways and subsequent treatment management approaches for children and young people can be highly variable, leading to diverse patient and clinical experiences. This study explores the DGBI diagnostic experiences of children and their families and the perspectives of clinicians in the New Zealand context.

**Methods:** Semi-structured interviews were conducted with 12 children with gastroduodenal DGBIs and their families and clinicians who care for children with DGBIs. Interviews were recorded, transcribed, and narratively analysed.

**Results:** Five children and family themes emerged: 1) how it all started, 2) the impacts symptoms had on child and family life, 3) their experiences with testing and investigations, 4) the perceptions and impacts of challenging clinical relationships, and 5) the uncertainness of trial and error treatments. Clinicians also identified five key themes: 1) navigating the complexity of presenting symptomology, 2) the challenging diagnostic investigation decision-making process, 3) navigating management and treatment approaches, 4) a lack of standardised clinical pathways, and 5) establishing therapeutic relationships with patients and families.

**Conclusion:** Children, their families, and clinicians confirmed the clinical complexity of DGBIs, the challenges of diagnosis and management, and the stress this places on therapeutic relationships. Clearer diagnostic pathways and new investigations that could provide improved identification and discrimination of DGBIs are needed to minimise the treat-test repeat cycle of care and improve health outcomes.

## BACKGROUND

Gastroduodenal symptoms such as nausea, vomiting, upper abdominal pain, bloating, early satiety, and reflux are commonly reported symptoms experienced by children with gastroduodenal disorders of gut-brain interaction (DGBIs). These symptoms can lead to significant discomfort and distress for children^1–5^, including poor health-related quality of life^6, 7^ sleep^8, 9^, physical activity, school, and social activities^10, 11^, and higher anxiety and depression.^12^ Additionally, these poorer outcomes have been found to persist into adulthood.^13^

Families with children with a DGBI often report poorer family functioning and higher stress levels than families with healthy children.^14, 15^ Furthermore, the financial burden can also be significant for the families themselves^16^ and the health sector. Enhancing diagnostic capabilities for DGBIs could contribute to increased patient satisfaction, better clinical outcomes, and a more comprehensive understanding of these often complex and multifaceted disorders.

Until the underlying pathophysiological mechanisms are better understood and validated biomarkers are established, the Rome Pediatric DGBI classification framework remains a well-established tool clinicians can utilise to make a positive diagnosis with minimal investigations.^17^ In addition to the Rome framework, various international Paediatric Societies have produced consensus-based condition-specific DGBI recommendations that reflect their healthcare jurisdictions and available diagnostic, management, and treatment options.

However, adherence and clinical utility remain variable despite these guidelines and recommendations.^18, 19^ It is clear from the international evidence that the diagnostic pathways and subsequent treatment management approaches for children and young people with gastroduodenal DGBIs can be highly variable.^14, 20, 21^ Research in New Zealand has reported a similar prevalence of DGBIs, including gastroduodenal DGBIs in children aged 4-18 years^12^, as global epidemiological studies. ^22, 23^ However, no studies have explored the diagnostic and treatment experiences of children with gastroduodenal DGBIs in New Zealand. Therefore, this study aims to gather the diagnosis and treatment journeys and experiences of New Zealand children, young people, and their families, as well as the experiences and perspectives of the health professionals who diagnose and care for children with gastroduodenal DGBIs.

## METHODS

A prospective qualitative study with a snowball recruitment methodology was adopted to gather rich and contextual data on the participants’ experiences.

This study received ethical approval from the Auckland Health Research Ethics Committee, New Zealand (AH22600) and is reported per the Standards for Reporting Qualitative Research (SRQR) (Table 4).^24^

### Participants

Children and young people (hereafter called children) and their families were recruited from clinical networks, social media posts, and online support groups. Children were included if they were aged between 5-18 years, lived in New Zealand, spoke English, and were diagnosed with a gastroduodenal DGBI. Gastroduodenal DGBIs were determined by each child meeting the criteria of at least one Rome IV Pediatric Functional Gastrointestinal Disorders Disorders of Gut-Brain Interaction categories - H1. Functional Nausea and Vomiting Disorders, and H2.

Functional Abdominal Pain Disorders, excluding H2c: IBS.^25^ A "family" included at least one legal guardian or parent of a child participant aged 5-16 years. However, it could include the whole family unit, including all legal guardians or parents, other significant family members, and siblings. Participants aged 17 or 18 could consent to participate in the interview independently if they preferred.

Health professionals such as family practitioners, paediatricians, paediatric gastroenterologists, clinical nurse specialists, and psychologists who diagnose and care for children with gastroduodenal DGBIs as part of their clinical roles were recruited through networks. No participating clinician was the current healthcare provider of any child participants, and the researchers had no prior relationship with any participants.

All participants took part in an online semi-structured virtual interview. Written informed consent was obtained from each participant. Separate semi-structured interview schedules were developed to guide each interview and enable flexibility in exploring unexpected issues that emerged during the interviews. The interviews with children and families explored narratives and experiences of the onset of symptoms, self-help seeking solutions, when health professional help was sought and why, and their experience of the health care journey. The interviews with the clinicians focused on exploring their experiences and challenges of diagnosing, managing treatment, and caring for children with DGBIs. Interviews were audio recorded, transcribed, de-identified, and coded using NVivo. Where names are used in quotes, they are fabricated to preserve participant confidentiality. Data saturation was reached with the 12^th^ child and family group and six clinicians.

There was no relationship between the researchers undertaking the research or analysing the data and the participants.

## ANALYSIS

A narrative analysis was followed.^26^ Each interview was read several times, and the text was parsed into meaningful items according to the purpose of the study. Each text item was categorised and coded to enable differences and similarities to be explored and to group into main themes.

## RESULTS

### Demographics

Twelve children were interviewed, with demographics provided in Table 1. Nine were interviewed with their mother, one interview included both mother and father, and two children were interviewed independently (both female). All children met the Rome IV criteria for a gastroduodenal DGBI, with four having overlapping symptoms with functional constipation (FC) (Table 1). However, they all reported that their upper gastrointestinal symptoms were the most bothersome.

**Table 1.** Demographics of participant children.

| Characteristics |  | Children<br>(n=12) |
| --- | --- | --- |
| Age (years), median (range) |  | 15.5 (12-18) |
| Female†, n (%) |  | 7 (58%) |
| Ethnicity | NZ European | 8 |
|  | Pasifika | 1 |
|  | Indian | 1 |
|  | South-East Asian | 1 |
|  | European | 1 |
| Rome IV | FD alone | 4 |
|  | Rumination | 1 |
|  | FN & FV | 2 |
|  | FAP-NOS | 1 |
|  | FD & FC | 2 |
|  | FN, FV & FC | 1 |
|  | FAP-NOS & FC | 1 |
| Children Interviewed with Parent |  | 10 |
| Children Interviewed Independently |  | 2 |

† Participants self-identified as male or female with no participant identified as gender diverse; FD= Functional Dyspepsia; FN= Functional Nausea; FV= Functional Vomiting; FAP-NOS= Functional Abdominal Pain not otherwise specified; FC= Functional Constipation

Six clinicians were interviewed (professions were general practitioner (GP), paediatrician, psychologist, nurse specialist, and paediatric gastroenterologist). The clinicians were from different regions and health settings across New Zealand. All worked full- or part-time in the public health sector.

The children and family interviews lasted between 45-110 minutes, and the clinician interviews lasted between 40-65 minutes.

## CHILDREN AND FAMILY THEMES

Five themes with a number of sub-themes (bolded in the text) were identified from the interviews with the children and their families. Table 2 presents the hierarchy of themes and example quotes from participants.

**Table 2.** Themes and quotes from interviews with Children and their Families.

| Children and their Families |  |  |
| --- | --- | --- |
| Theme | Sub-theme | Direct Quotes |
| **1. How it all started** | Acute presentation | ...early Sunday he woke up really early in real distress, crying, really complaining that his tummy really hurt. So off to ED [emergency department] we went. They took some bloods and asked about bowels... Poked and prodded and took an x-ray. All normal. That surprised me. But in some ways I thought ok, that's good... So we went home. (P 10) |
|  | Slow burner | ...complaining of tummy pain and nausea for a couple of months off and on I was a bit worried that it hadn't gone away. (P 07) |
| **2. Impact on Life** | School attendance and learning | So many days off school this past year. I am worried that she will give up because she is behind in everything. (P 05)<br>I home-school him now, and we manage the ups and downs without a lot of stress. (P 12) |
|  | Psychological impacts | It gets lonely... I want to join in, but I am worried that I will feel awful, so I don't. (P 01) |
|  | Relationships with friends | I miss them. They don't really invite me to anything any more... (P 02)<br>It is hard sometimes with my friends, they are really supportive but it's hard for them, I think, to know why I don't just eat. (P 06) |
|  | Stressful family dynamics | I know she gets a bit sad at times. Especially when her friends are all getting out and doing things. It is heartbreaking for me. (P 06)<br>...can get a bit heated amongst them... we can't do some things ... so he feels he gets no attention... can't do the things he wants to do (P 09)<br>I tried to get him to eat more when we all sit down, you know, for dinner, but it ends up being disastrous, so lots of options and no pressure seems to be working. (P 04) |
|  | Financial impact | It takes a bit of a hit, income-wise... I stopped work, but I needed to, so I could help home-school... (P 05) |
|  | Avoiding or restricting food | Food is a nightmare. I used to love it and loved all sorts of flavours. Now, I feel it is a bit, umm, you know, will I be ok eating this or not? (P 02) |
|  | Coping strategies/initiatives | We break up the day... short intensity... then rest... it works (P 05)<br>I stop and focus and do this deep breathing (P 01) |
| 3. Investigations, Normal outcome, and a Diagnosis | Access to Specialist servoces | We have been to the GP 3-4 times over the past 6-12 months. Apart from blood and poo tests at the beginning, which were normal, he hasn't done any more tests. Yet nothing has improved... (P 08)<br>... we just don't meet the criteria to see a specialist. (P 03) |
|  | Normal investigation outcomes | The first time at ED [emergency department]...we had a few tests, x-rays, ultrasound and blood. All normal ... a relief really...(P 07)<br>Being told the tests results, all of them, were all normal, just surprised me.... Deep down I thought that they must be wrong. (P 09) |
|  | New diagnostic tools | ... been told that the current tests are not going to tell us anything...you know, they will all be normal... be nice to have something that will tell us what is wrong ... (P 08) |
|  | Confirming a DGBI diagnosis | It must have been a year after all this started that we first heard the term functional disorders. She has this dyspepsia diagnosis... (P 05)<br>Hearing about these gut-brain disorders... functional nausea...first made me think they thought it was all in her head. I was ready to fight for her, I mean she was really unwell... I have done a lot of reading about these now... so we are functional nausea and vomiting disorder ... [laugh] it just doesn't feel like a diagnosis... (P 03) |
| 4. Clinical Relationships | Disempowered | ... those subtle interactions ... like they don't believe it is real... that **disempower** me. (P 04) |
|  | Greater sense of control | It took a while, but I now have a really great relationship with the docs. They really listened to me. It made me feel that I can manage this stuff. (P 01) |
| 5. Trial and Error Treatment | Trial and error | We've been given diets, medicines, diets and medicines, plus we have done our own stuff ... acupuncture, prebiotics, probiotics. Some seem to work for a bit, then stop. It makes her life a bit stressful. I know she thinks, how long will this work until it doesn't? Then back to the hospital for the next round... (P 09)<br>We have laughed a few times about the medicine lotto. Will we win this time or will it be "not a winning medicine" [laughter]. (P 02)<br>...feel it is all a big experiment. No one really knows what to do or what will work. (Fam 07) |
|  | Pass the parcel | I was sort of passed out of the GI services and on to psychological people and to GP then back again. **(P 01)** |

### Theme 1. How it all started

The first theme explored the reflections children and their families had on how it all started as a way to contextualise where they are now in their journey. Two main starting points emerged, with the two groups having differential experiences with care. Firstly, an **acute** start was described, where the child had an unexpected episode of abdominal pain or vomiting that was unrelenting and resulted in families seeking medical care at the emergency department (ED) or urgent care centre. The second was the **slow burner** start, where, over time, there was an accumulation of events, behaviours, and experiences that signalled something was wrong and thus sought medical advice, primarily from their GP. Participants were equally distributed between these two starting points.

All participants recalled a comprehensive physical examination and laboratory tests during their first interaction with medical care. For the acute group, two children had abdominal X-rays, and one had an abdominal ultrasound. In all cases, participants reported that the investigation results were normal. This elicited a sense of relief for the acute group. In contrast, the slow burner group reported feeling somewhat unsatisfied with the lack of conclusive results to explain their child’s symptoms.

After the initial medical care visit, the acute group commented that they were given little advice on managing the symptoms, including seeing their GP if symptoms persist or worsen. In contrast, the slow burner group reported receiving prescriptions for take-as-needed anti-emetics (n=2) and laxatives (n=3), with one participant receiving analgesia, anti-emetics, and laxatives. All reported being provided dietary advice but recalled that this was not structured guidance and was "*pretty vague"*.

### Theme 2. Impact on life

The third theme explored the impact of the symptoms on their lives. The parent participants frequently mentioned the impact of symptoms on **school attendance and learning**, with almost half of the children reporting being home-schooled or having periods of home-schooling due to their symptomatology. Their symptoms also resulted in negative **psychological impacts** for the child and family, including anxiety and depression. **Relationships** were also negatively impacted; for example, the children experienced the loss of school friendships.

Parents also mentioned the **stress** placed on the internal family dynamics, and two parents also mentioned the **financial impact.** This was particularly highlighted by participants who had to home-school their children.

All the children reported a shift in their relationship with food since the onset of their symptoms, including **food avoidance, restriction, or fear.**

It was evident that parents and the children employed various **coping strategies and initiatives** to lessen the impact of the symptoms on their lives. These coping strategies varied widely; for example, one family home-schooled their child, and they talked about breaking up the day with lots of breaks to accommodate times when symptoms were "*particularly bad*".

Another spoke about "*deep breathing and not focusing on the symptoms"*, whereas another said they "*went to bed until they felt better"*. These varying strategies reflected the diverse family dynamics and the specific symptoms experienced by the children.

### Theme 3. Investigations, the Normal Outcome, and the Diagnosis

The third theme highlighted the clinical care pathway experienced, with differential experiences noted across participants. Only two children were referred to specialist services at their first medical presentation. Three participants reported that the outcome of the first referral was that they *did not meet the threshold for a specialist appointment* and that care remained with the GP. One family decided to go privately. **Access to a specialist service** was reported to take 3-12 months and 4-5 visits to the GP before being referred.

Once children were under specialist care, more invasive tests such as endoscopies (n=8), radiological tests (CT scans, n=2), and nuclear tests (scintigraphy, n=3) were undertaken. No one reported a clinically significant outcome from the range of tests, and the continued surprise at the **normal test result outcomes** was a common theme. The lack of correlation between the intensity of symptoms and a tangible investigation outcome was incomprehensible for some families. For others, the normal results were viewed as *a relief*.

Most of the participants described current diagnostic tools and processes as limited. They remarked that the current tools did not provide enough insights and that there was a need for **new diagnostic tools** that could offer more precise and efficient methods to identify their children’s disorders. This would facilitate earlier and more accurate diagnoses that could enhance the clinical experience by alleviating the emotional and physical burden of ongoing symptoms. Plus, by providing a clearer diagnosis, the doctors can tailor treatment plans from the outset, thus reducing the treat-test-treat-repeat cycle.

Despite numerous interactions with primary care, many participants reported that the term "functional" was not introduced as a potential **diagnosis** for their child’s symptoms until they accessed specialist services. Some participants recalled that the term "functional" was **not** well-explained, leaving them feeling that their concerns were not taken seriously. These participants were more inclined to seek out additional medical opinions.

### Theme 4. Clinical Relationships

The fourth theme explored the participants’ perceptions of their relationships with healthcare providers. A few participants’ perceptions of the early negative encounters with the health sector profoundly influenced subsequent interactions, often setting a challenging and combative tone with the clinical relationship. Some participants highlighted that they had to persistently advocate for their child’s condition to be taken seriously or to push for further testing, which contributed to higher levels of dissatisfaction with the care provided and feelings of **disempowerment**.

In contrast, participants who experienced a positive relationship and open communication with their clinical team felt supported and regarded themselves as active partners in the decision-making process. One participant recounted how her clinical team offered support by attentively listening to and valuing her opinions and actively encouraging her to monitor her progress with different treatments. This approach gave her a **greater sense of control** over her symptoms and significantly boosted her confidence in managing them.

### Theme 5. Trial and Error Treatment Experience

The final theme explored participants’ treatment and management experiences, who often described a **trial-and-error approach** to their care. They recognized that, due to the absence of an identifiable physical problem, treatment strategies focused on experimenting with different dietary modifications and medications to alleviate symptoms. Participants expressed frustration with how time-consuming and challenging this approach could be, especially when they were eager for immediate symptom relief.

Nevertheless, most of the participants also remarked that they understood that this method aimed to find the right treatment balance without excessive reliance on medications.

However, the trial-and-error approach also had its drawbacks. One teenage participant likened her experience to being a parcel in a children’s party game of **pass-the-parcel**, illustrating how she felt shuffled between clinicians, given different explanations and medicines, without clear communication concerning what to expect. This analogy underscored the difficulties patients faced in navigating the healthcare system and the need for a more cohesive and coordinated approach to care.

Participants frequently highlighted the lack of timely access to support services such as psychologists or dietitians, which exacerbated the challenges they faced in managing their symptoms. The lack of access often left families feeling unsupported and overwhelmed as they tried to navigate complex treatment regimens on their own. This contributed to feelings of frustration and helplessness and, for some, also eroded their relationship with the clinicians caring for them.

## CLINICIAN THEMES

Five key themes, with a number of sub-themes (bolded in the text), emerged from the clinician interviews. Table 3 presents the hierarchy of clinician themes, with example quotations.

**Table 3.**
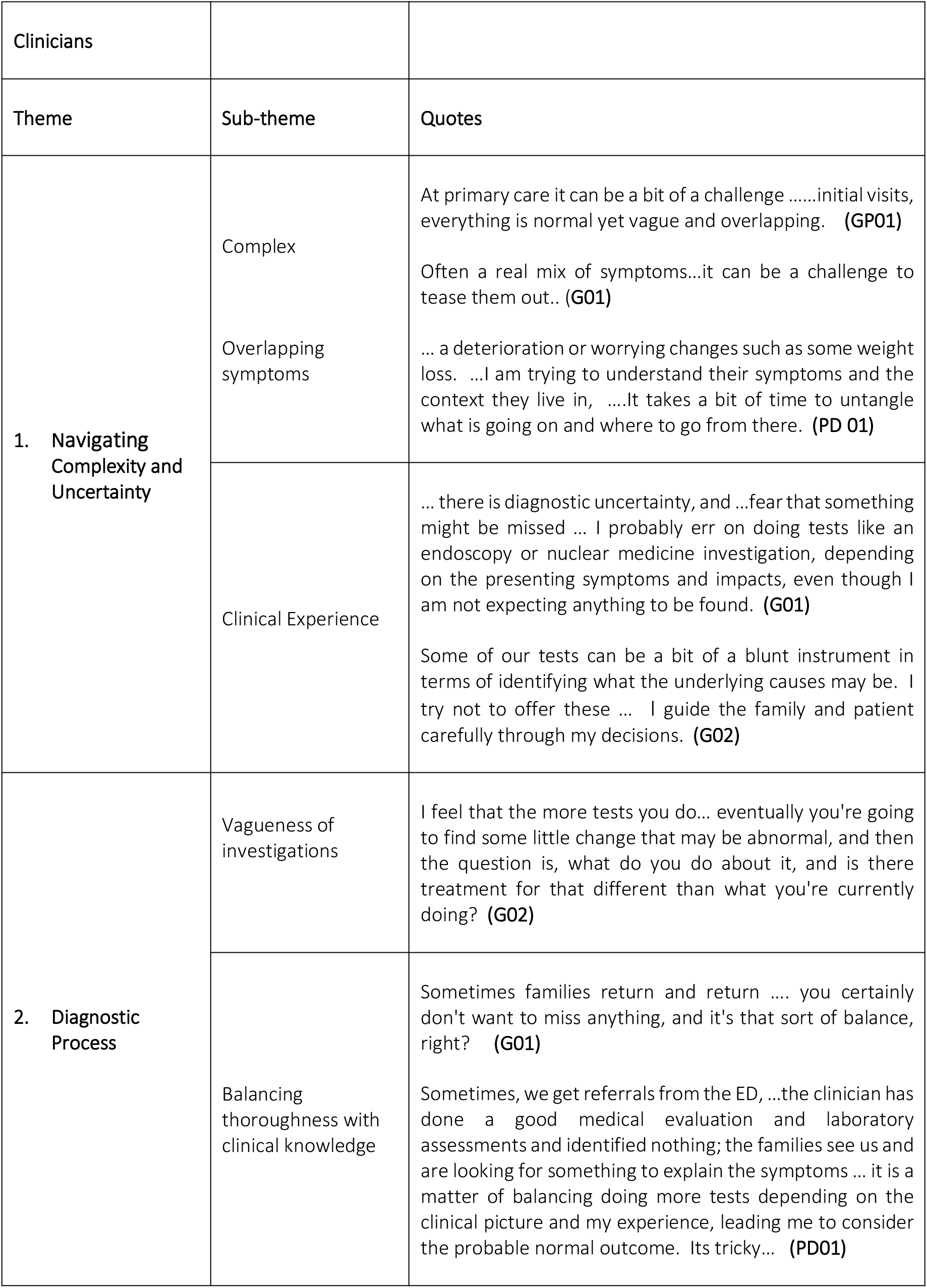
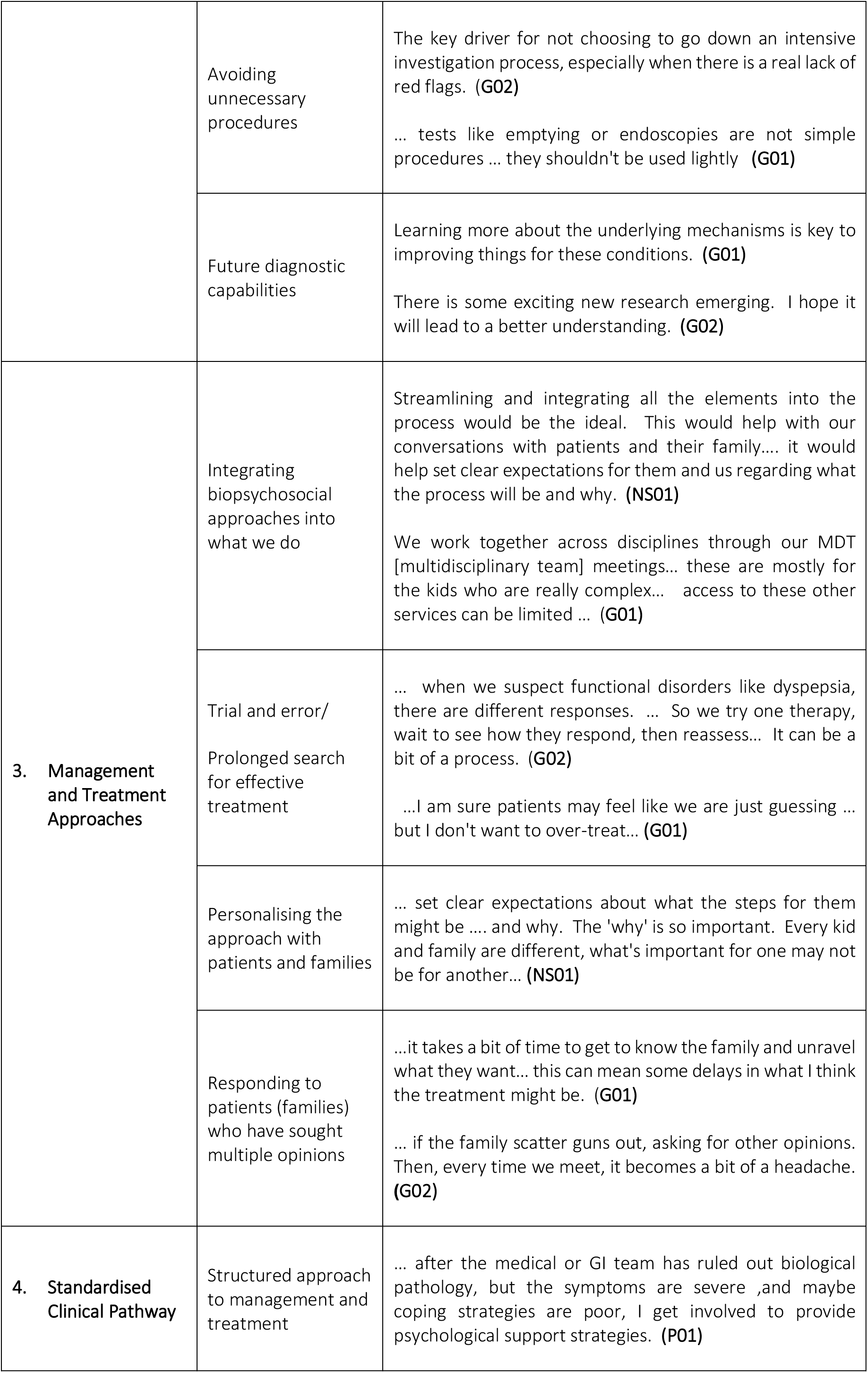
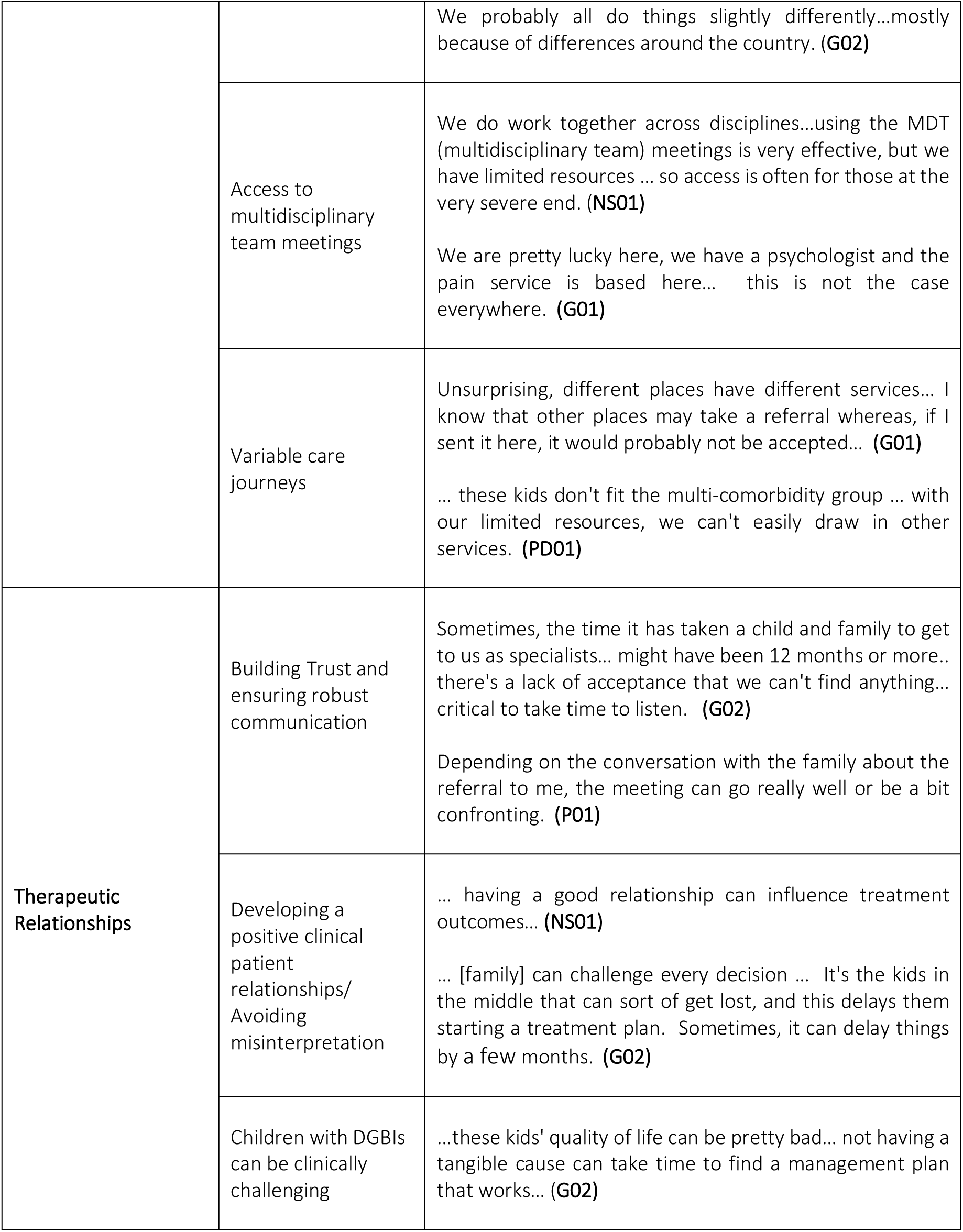
Themes and quotes from interviews with clinicians who care for children with DGBIs as part of their clinical role.

**Table 4.**
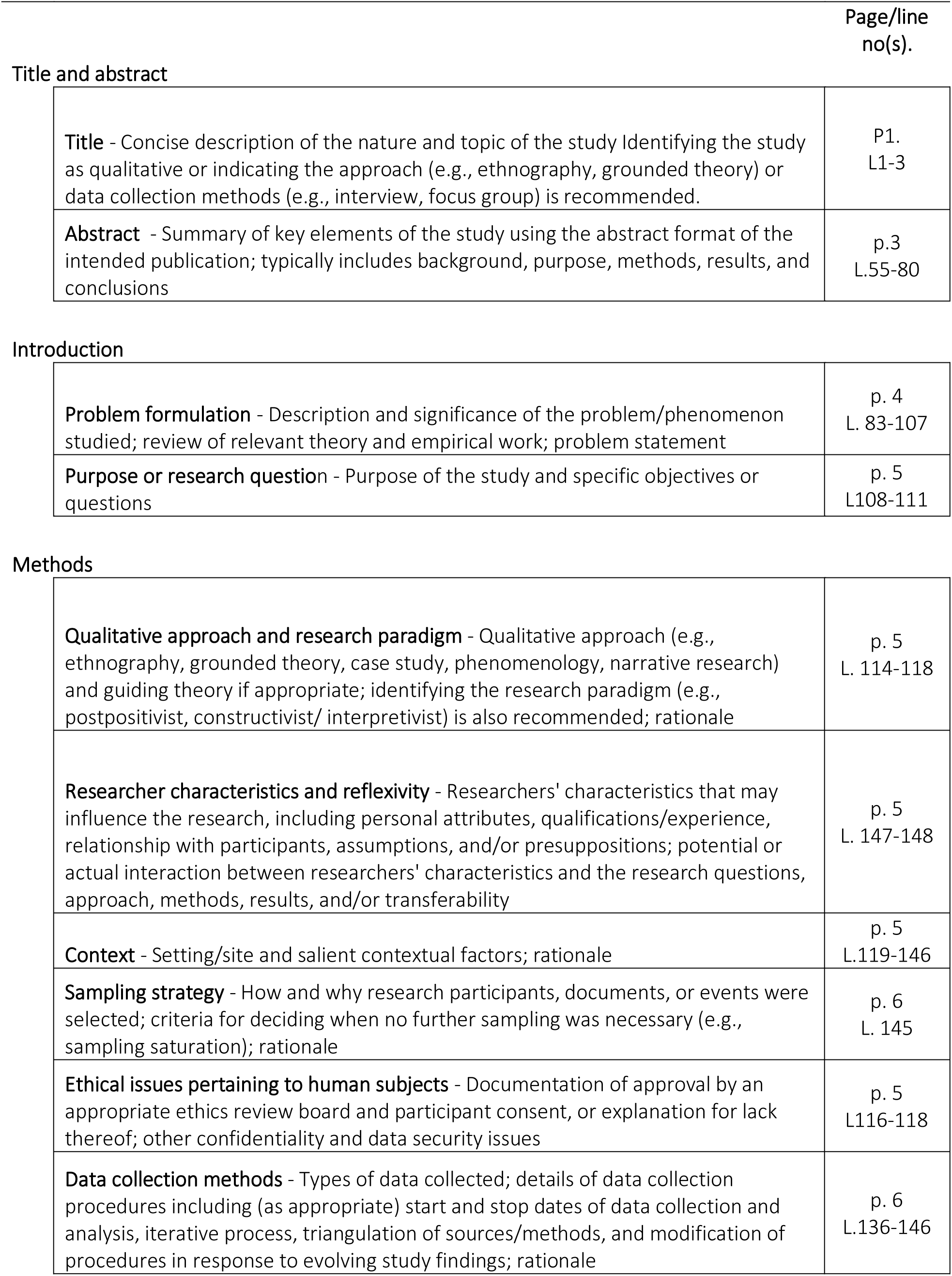
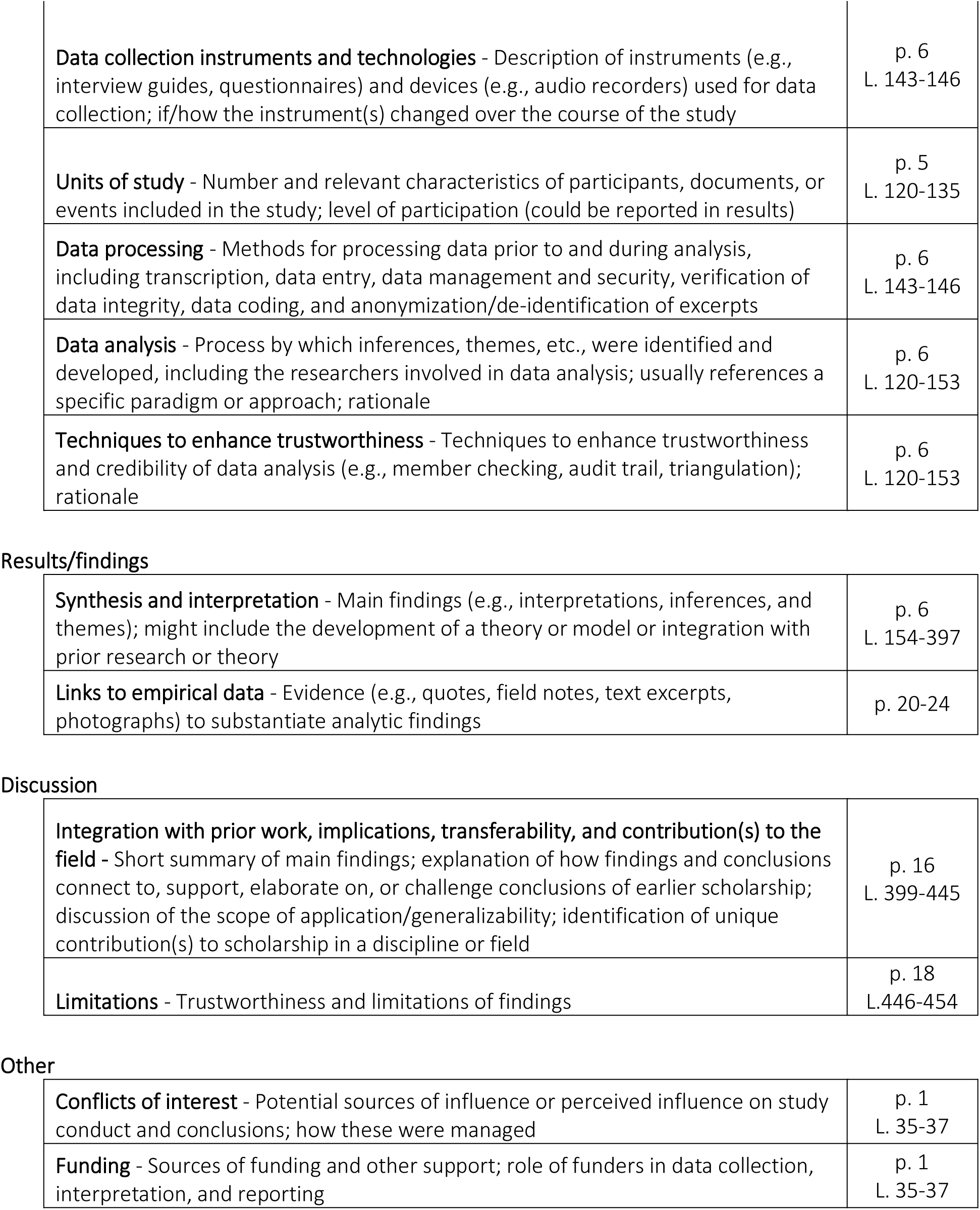
Standards for Reporting Qualitative Research (SRQR)

|  | Page/line no(s). |
| --- | --- |
| **Title and abstract** |  |
| **Title** - Concise description of the nature and topic of the study Identifying the study as qualitative or indicating the approach (e.g., ethnography, grounded theory) or data collection methods (e.g., interview, focus group) is recommended. | P1.<br>L1-3 |
| **Abstract** - Summary of key elements of the study using the abstract format of the intended publication; typically includes background, purpose, methods, results, and conclusions | p.3<br>L.55-80 |
| **Introduction** |  |
| **Problem formulation** - Description and significance of the problem/phenomenon studied; review of relevant theory and empirical work; problem statement | p. 4<br>L. 83-107 |
| **Purpose or research question** - Purpose of the study and specific objectives or questions | p. 5<br>L108-111 |
| **Methods** |  |
| **Qualitative approach and research paradigm** - Qualitative approach (e.g., ethnography, grounded theory, case study, phenomenology, narrative research) and guiding theory if appropriate; identifying the research paradigm (e.g., postpositivist, constructivist/ interpretivist) is also recommended; rationale | p. 5<br>L. 114-118 |
| **Researcher characteristics and reflexivity** - Researchers' characteristics that may influence the research, including personal attributes, qualifications/experience, relationship with participants, assumptions, and/or presuppositions; potential or actual interaction between researchers' characteristics and the research questions, approach, methods, results, and/or transferability | p. 5<br>L. 147-148 |
| **Context** - Setting/site and salient contextual factors; rationale | p. 5<br>L.119-146 |
| **Sampling strategy** - How and why research participants, documents, or events were selected; criteria for deciding when no further sampling was necessary (e.g., sampling saturation); rationale | p. 6<br>L. 145 |
| **Ethical issues pertaining to human subjects** - Documentation of approval by an appropriate ethics review board and participant consent, or explanation for lack thereof; other confidentiality and data security issues | p. 5<br>L116-118 |
| **Data collection methods** - Types of data collected; details of data collection procedures including (as appropriate) start and stop dates of data collection and analysis, iterative process, triangulation of sources/methods, and modification of procedures in response to evolving study findings; rationale | p. 6<br>L.136-146 |
| **Data collection instruments and technologies** - Description of instruments (e.g., interview guides, questionnaires) and devices (e.g., audio recorders) used for data collection; if/how the instrument(s) changed over the course of the study | p. 6<br>L. 143-146 |
| **Units of study** - Number and relevant characteristics of participants, documents, or events included in the study; level of participation (could be reported in results) | p. 5<br>L. 120-135 |
| **Data processing** - Methods for processing data prior to and during analysis, including transcription, data entry, data management and security, verification of data integrity, data coding, and anonymization/de-identification of excerpts | p. 6<br>L. 143-146 |
| **Data analysis** - Process by which inferences, themes, etc., were identified and developed, including the researchers involved in data analysis; usually references a specific paradigm or approach; rationale | p. 6<br>L. 120-153 |
| **Techniques to enhance trustworthiness** - Techniques to enhance trustworthiness and credibility of data analysis (e.g., member checking, audit trail, triangulation); rationale | p. 6<br>L. 120-153 |
| Results/findings |  |
| **Synthesis and interpretation** - Main findings (e.g., interpretations, inferences, and themes); might include the development of a theory or model or integration with prior research or theory | p. 6<br>L. 154-397 |
| **Links to empirical data** - Evidence (e.g., quotes, field notes, text excerpts, photographs) to substantiate analytic findings | p. 20-24 |
| **Integration with prior work, implications, transferability, and contribution(s) to the field** - Short summary of main findings; explanation of how findings and conclusions connect to, support, elaborate on, or challenge conclusions of earlier scholarship; discussion of the scope of application/generalizability; identification of unique contribution(s) to scholarship in a discipline or field | p. 16<br>L. 399-445 |
| **Limitations** - Trustworthiness and limitations of findings | p. 18<br>L. 446-454 |
| Other |  |
| **Conflicts of interest** - Potential sources of influence or perceived influence on study conduct and conclusions; how these were managed | p. 1<br>L. 35-37 |
| **Funding** - Sources of funding and other support; role of funders in data collection, interpretation, and reporting | p. 1<br>L. 35-37 |

### Theme 1. Navigating Complexity and Uncertainty

The first clinician theme explored how managing DGBIs involves navigating **complex symptoms** and an incomplete understanding of underlying mechanisms. As clinicians, this meant these symptoms required a nuanced approach that appreciates the diversity of symptoms and their impact on patients’ lives.

Depending on where they were in the clinical pathway (primary care to the specialist) and their confidence to make a functional diagnosis, clinicians responded differently to the lack of a clear aetiology. The Rome criteria were well known but were reported as difficult to apply in practice (I would add why here). Different international and local guidelines were also mentioned; however, patients presented with a mix of **dominant overlapping symptoms**, which made applying either Rome or the guidelines difficult. As such, there was a tendency to rely more on **clinical experience** to suit a patient’s unique symptomatology and needs.

### Theme 2. Diagnostic Process

The second theme explored the current diagnostic decisions and processes clinicians implemented and how these were highly correlated with the complexity and uncertainty of these symptoms. The current **diagnostic tools were viewed as vague** in their ability to provide a clear diagnostic outcome in most circumstances, with ’normal’ or ’nothing abnormal’ outcomes reported as somewhat dissatisfying.

When selecting investigations, the focus was on carefully **balancing thoroughness** and potential benefits with the necessity of **avoiding unnecessary procedures** with low yields and minimising the burden of tests. This approach was especially important in fostering a strong therapeutic relationship, as many patients may have waited over 12 months for a specialist appointment and were highly anxious about what may be wrong. However, it was also important to ensure that patients did not end up on an "investigative roundabout", which often shifted the focus from management and treatment plans to searching for an elusive diagnosis . Effective and empathetic communication was reported as crucial for streamlining the clinical care pathway process.

Despite the frustration currently experienced, there was optimism for **future diagnostic capabilities** that would provide some certainty to the underlying aetiology of DGBIs, which was somewhat missing with the current diagnostic investigations. Clinicians remarked that the current investigations played a role in ensuring there was no potential organic condition, inflammation, or other co-existing structural issues. However, despite conducting these tests, they remarked that they were pretty confident that the outcomes for many of these children would likely be normal. Many highlighted that the new research into the gut microbiome, metabolomics, and innovations in non-invasive imaging technology held promise for more accurate and earlier identification of the underlying mechanisms of DGBIs. The desired outcome highlighted was that these new investigations would help direct decisions for those needing further investigation and also open up new insights into personalised treatments and ultimately improve the lives of children and their families.

### Theme 3. Management and Treatment Approaches

The third clinician theme discussed how DGBIs present significant treatment and management challenges due to their complex and heterogeneous nature. Effective management requires a comprehensive **biopsychosocial approach,** integrating multiple elements such as lifestyle and dietary modifications, pharmacological treatments, and psychological therapies. However, treatment for DGBIs continues to be a matter of **trial and error** for patients because symptoms vary widely.

Clinicians noted how trial-and-error treatment approaches were not unique, with many other chronic conditions relying on a **personalised approach to** selecting different therapy combinations to find what combination alleviates the patient’s symptoms. However, without a clear understanding of the mechanisms involved for many DGBIs, the trial and error approach can lead to a prolonged search for effective treatments, and patients and families often endure significant emotional distress and frustration during this process.

The distress and frustration experienced by families were reported as a common reason some families sought **multiple opinions** from different medical professionals, which sometimes led to further delays in care or treatments that are conflicting without realising. Irrespective of the reasons for treatment delays, participants remarked that delays in confirming a diagnosis were one of the main reasons for exacerbating or worsening a child’s symptoms.

### Theme 4. Standardised Clinical Pathway

The fourth clinician theme highlighted how a clinical pathway offers a **structured approach to management and treatment** that integrates evidence-based practices. Participants responded that there was no clear standardised pathway to join different healthcare services. Different regions across New Zealand had different referral pathways from GP to hospital/specialist care. Differences emerged in the thresholds for sending a referral, criteria at hospital/specialist services accepting a referral, the internal triaging process at the specialist clinic (which determines wait times), and the individual clinician variation in their clinical care approaches.

There were also notable variations in **access to multidisciplinary services** like psychologists or dietitians, which were considered essential to management. In many cases, these services had limited staff numbers and long wait lists, and in some geographic locations, these services were unavailable. All participants highlighted these challenges as reasons their patients had such **variable clinical journeys.**

### Theme 5. Therapeutic Relationships

The final theme highlighted that the therapeutic relationship forged between the doctor, patient, and family was a cornerstone of health care. **Building trust and ensuring robust communication** was paramount, as they enhanced patient/family engagement as partners in the process, positively impacting the management. Given the often misunderstood and prolonged nature of these conditions, it was important that patients and their families feel listened to, validated and supported. Establishing a **positive clinical-patient (family) relationship** was highlighted as crucial in empowering patients. Through this relationship, valuable knowledge is shared, leading to shared decisions that enable patients to gain the confidence needed to manage their conditions effectively. This empowerment leads to better management of their health and, ultimately, an improved quality of life.

In contrast, participants highlighted many barriers to establishing and maintaining a positive therapeutic relationship. Barriers such as parents’ desire for a curative outcome, which was only possible with a ’proper’ diagnosis, led to confrontational interactions that were challenging to work through. The difficulty in explaining DGBIs and the general lack of clarity of the underlying mechanisms sometimes created parental perceptions that their concerns were not adequately addressed. Finally, discussing the psychological factors that influence DGBIs, clinicians reported that, at times, families had **misinterpreted** or **misunderstood** these conversations as their child’s concerns were being overlooked or dismissed. Taking time to have these interactions is important, but this was not always possible in busy healthcare settings.

Furthermore, access to other services, such as psychological and social support, was ’..*non-existent’, ’..limited’* or ’..*had long wait times’.* The clinicians reflected that many of the issues and barriers experienced were a symptom of a lack of a well-staffed multidisciplinary team to support the diagnostic and treatment pathway. The overarching sentiment from the clinicians was that children with DGBIs (and their families) were some of the **most clinically challenging**.

## DISCUSSION

Children, family, and clinician participants acknowledged that diagnosing DGBIs is complex due to the variability and severity of symptom presentation and the impact on quality of life. For example, a recurring theme for the children and families was the lengthy and often convoluted clinical journey to achieve what was, for some, an unsatisfactory diagnosis. While the type, timing, and frequency of investigations varied, a common outcome was the absence of abnormal findings. For clinicians, this absence of abnormalities provided the certainty needed to make a DGBI diagnosis; however, for families, it sometimes resulted in confrontational interactions and a decline in the therapeutic relationship.

The desire for diagnostic certainty was similarly strong between groups; however, there was a notable incongruence in how clinicians and families understood certainty. While clinicians often viewed certainty as the ability to rule out serious conditions and, where possible, provide a probable diagnosis based on symptoms as per Rome criteria^25^, families tended to seek more definitive answers, often hoping for clear-cut diagnoses with identifiable causes and straightforward treatment plans. This mismatch in expectations led to frustration and dissatisfaction and resulted in some families seeking multiple medical opinions. This cycle of behaviour has been found to lead to delays in treatment initiation, poorer outcomes and higher healthcare costs.^27, 28^ These experiences were recounted as contributing to a somewhat combative clinician-patient/family therapeutic relationship. Similar findings are reported in the international literature.^29–32^

Furthermore, clinicians and families acknowledged the limitations of the investigations currently available and that new tests are needed that offer more nuanced indicators and actionable biomarkers. The lack of understanding of aetiopathogenesis can result in similar symptoms in patients that are not always ameliorated by similar treatments, resulting in a trial-and-error approach to treatment. ^33^ For the children and the families, reducing this cycle through more effective diagnostic technologies would reduce the diagnostic journey, provide more certainty of the care pathway, and positively impact their children’s quality of life. For clinicians, feeling more confident with the diagnosis and management plan could improve the clinician-patient relationship. In addition, introducing the term functional or DGBI early in the diagnostic journey is essential and has been found to help avoid over-investigation.^34^

Research reports that communication is an essential skill for clinicians caring for patients with DGBIs.^35–37^ This study’s findings illustrate that, at times, clinician-patient/family communication was difficult and impacted the therapeutic relationships, which, in turn, caused dissatisfaction with what was perceived as a lack of a care pathway. Other research reflects similar findings and highlights the importance of a positive therapeutic relationship in that it promotes relational continuity and positively impacts patient health outcomes.^14, 20, 21^

Research evidence also highlights the effectiveness of a multidisciplinary team involved in caring for and managing children (and families) with DGBIs.^38–40^ Effectiveness is shown through clearer care pathways, shorter diagnostic journeys, and early initiation of treatment. The participants’ conversations in this research revealed a distinct lack of a multidisciplinary approach to care and management due to the limited availability of dieticians and psychologists and variable access to these services in New Zealand. Improving the availability and access to multidisciplinary care was highlighted as a crucial area needing remediation.

Such enhancements would ensure patients benefit from a comprehensive approach that integrates medical, dietary, psychological, and other specialized support, offering a more holistic and effective management strategy for conditions like DGBIs. This approach supports better patient outcomes and addresses the diverse needs of patients and their families, thereby improving overall satisfaction with healthcare delivery.^50^

While data saturation was achieved in this study, some limitations are worth noting. Patient participants were predominantly New Zealand European, with one Pasifika and no Māori participants [Indigenous population of New Zealand). However, Māori and Pasifika are represented in the prevalence of DGBIs in a cohort of New Zealand children.^12^ As food and meals in Pasifika and Māori cultures are embedded within many cultural interactions^41^, more research into the impact of DGBIs and the diagnostic journeys for Māori and Pasifika children and families is an important area for future research. Additionally, this research did not gather details of all the investigations, therapies, and healthcare visits or the participants. Reviewing these details alongside these narratives can help provide a better understanding of the number of health services contacts and enable the costs of the current clinical care approach to be calculated.

## Conclusion

This study explored the diagnostic and treatment experiences of children with DGBIs and their families and clinicians who care for patients with DGBIs in a New Zealand setting. The findings highlighted a significant impact of gastroduodenal symptoms on the quality of life of these children. Inadequacies in the current diagnostic investigations and the overall vagueness associated with being given and giving a DGBI diagnosis were highlighted throughout the interviews. The reliance on a trial-and-error management approach further contributed to dissatisfaction with the experience among children, their families, and clinicians. While a multidisciplinary model of care was recommended, introducing new investigations that could provide actionable biomarkers was highly desirable. If used early in the diagnostic and management pathway, these tools could support earlier diagnoses, enable personalized treatment plans, and help identify patients who would most benefit from the current investigation tools.

## Acknowledgements and Funding

The authors would like to thank the children, their families and the clinicians who took part in this research. This study was supported by the Health Research Council of New Zealand.

## Author Contributions

GH, CK, and GOG were involved in the conception and design of the study. GH undertook the data collection and analyses and wrote the first draft. All authors contributed to critical revisions and final approval of the manuscript.

## Data Availability

The data supporting the findings in this manuscript are available through the corresponding 50 author (GH) upon reasonable request and ethical approval.

## References

1. Calvano C, Warschburger P. Chronic Abdominal Pain in Children and Adolescents: Parental Threat Perception Plays a Major Role in Seeking Medical Consultations. Pain research & management 2016;2016:1–10.

2. Warschburger P, Hänig J, Friedt M, et al. Health-related quality of life in children with abdominal pain due to functional or organic gastrointestinal disorders. Journal of Pediatric Psychology 2014;39:45–54.

3. Park R, Mikami S, LeClair J, et al. Inpatient burden of childhood functional GI disorders in the USA: an analysis of national trends in the USA from 1997 to 2009. Neurogastroenterology & Motility 2015;27:684–692.

4. Robin SG, Keller C, Zwiener R, et al. Prevalence of Pediatric Functional Gastrointestinal Disorders Utilizing the Rome IV Criteria. The Journal of Pediatrics 2018;195:134–139.

5. Ansems SM, Ganzevoort IN, van Tol DG, et al. Qualitative study evaluating the expectations and experiences of Dutch parents of children with chronic gastrointestinal symptoms visiting their general practitioner. BMJ open 2023;13:e069429–e069429.

6. de Jesus CDF, de Assis Carvalho M, Machado NC. Impaired Health-Related Quality of Life in Brazilian Children with Chronic Abdominal Pain: A Cross-Sectional Study. Pediatric gastroenterology, hepatology & nutrition 2022;25:500–509.

7. Rajindrajith S, Zeevenhooven J, Devanarayana NM, et al. Functional abdominal pain disorders in children. Expert Review of Gastroenterology & Hepatology 2018;12:369–390.

8. Robbertz AS, Shneider C, Cohen LL, et al. Sleep Problems in Pediatric Disorders of Gut–Brain Interaction: A Systematic Review. Journal of pediatric psychology 2023;48:778–786.

9. Kim HJ. Importance of sleep quality in functional abdominal pain disorder in pediatric patients. Sleep and biological rhythms 2022;20:81–85.

10. Lewis ML, Palsson OS, Whitehead WE, et al. Prevalence of Functional Gastrointestinal Disorders in Children and Adolescents. The Journal of Pediatrics 2016;177:39–43.e3.

11. Brun R, Kuo B. Review: Functional dyspepsia. Therapeutic Advances in Gastroenterology 2010;3:145–164.

12. Vernon-Roberts A, Alexander I, Day AS. Prevalence of Functional Gastrointestinal Disorders (Rome IV Criteria) among a Cohort of New Zealand Children. Gastrointestinal Disorders 2023;5:261–272.

13. Jones MP, Koloski NA, Walker MM, et al. A Minority of Childhood Disorders of Gut-Brain Interaction Persist Into Adulthood: A Risk-Factor Analysis. Official journal of the American College of Gastroenterology | ACG 2024:10.14309/ajg.0000000000002751.

14. Feingold JH, Drossman DA. Deconstructing stigma as a barrier to treating DGBI: Lessons for clinicians. Neurogastroenterology and motility 2021;33:e14080-n/a.

15. Garr K. The impact of pediatric disorders of gut-brain interaction on the family: the roles of mental health and somatic symptoms. Department of Psychology. Volume Doctor of Philosophy: University fo Cincinnati, 2022.

16. Calvano C, Warschburger P. Quality of life among parents seeking treatment for their child’s functional abdominal pain. Quality of Life Research 2018;27:2557–2570.

17. Drossman DA. Functional Gastrointestinal Disorders: History, Pathophysiology, Clinical Features, and Rome IV. Gastroenterology 2016;150:1262–1279.e2.

18. Quitadamo P, Urbonas V, Papadopoulou A, et al. Do Pediatricians Apply the 2009 NASPGHAN–ESPGHAN Guidelines for the Diagnosis and Management of Gastroesophageal Reflux After Being Trained? Journal of pediatric gastroenterology and nutrition 2014;59:356–359.

19. Vieira SCF, Gurgel FM, Leão MZ, et al. Survey on the Adherence to the 2009 NASPGHAN-ESPGHAN Gastroesophageal Reflux Guidelines by Brazilian Paediatricians. Journal of pediatric gastroenterology and nutrition 2018;67:e1–e5.

20. Waheed A, Malone M, Samiullah S. Functional Gastrointestinal Disorders: Functional Gastrointestinal Disorders in Children. FP Essentials 2018;466:29–35.

21. Breen M, Murphy KP, O’Neill SB, et al. The utilisation and diagnostic yield of radiological imaging in a specialist functional GI disorder clinic: an 11-year retrospective study. European radiology 2014;24:3097–3104.

22. Vernon-Roberts A, Alexander I, Day AS. Systematic Review of Pediatric Functional Gastrointestinal Disorders (Rome IV Criteria). J Clin Med 2021;10.

23. Velasco C, Collazos-Saa L, García-Perdomo H. A systematic review and meta-analysis in schoolchildren and adolescents with functional gastrointestinal disorders according to rome iv criteria. Arquivos de Gastroenterologia 2022;59:304–313.

24. O’Brien BC, Harris IB, Beckman TJ, et al. Standards for Reporting Qualitative Research: A Synthesis of Recommendations. Academic Medicine 2014;89.

25. Douglas Drossman, Lin Chang, William Chey, et al. Rome IV Pediatric Functional Gastrointestinal Disorders – Disorders of Gut-Brain Interaction, First Edition Raleigh, North Carolina: Rome Foundation, 2016.

26. Thomas D. A General Inductive Approach for Analyzing Qualitative Evaluation Data. American Journal of Evaluation 2006;27:237–246.

27. Jia M, Lu P, Khoo J, et al. Delay in diagnosis is associated with decreased treatment effectiveness in children with rumination syndrome. Journal of pediatric gastroenterology and nutrition 2024.

28. Biernikiewicz M, Germain N, Toumi M. Second opinions, multiple physician appointments, and overlapping prescriptions in the paediatric population: A systematic literature review. Journal of evaluation in clinical practice 2020;26:1761–1767.

29. Gamboa HE, Sood MR. The Spectrum of Functional GI Disorders. Cham: Springer International Publishing, 2021:255-264.

30. Macpherson AK, Kramer MS, Ducharme FM, et al. Doctor shopping before and after a visit to a paediatric emergency department. Paediatr Child Health 2001;6:341–6.

31. Drossman DA, Chang L, Deutsch JK, et al. A Review of the Evidence and Recommendations on Communication Skills and the Patient -Provider Relationship: A Rome Foundation Working Team Report. Gastroenterology 2021;161:1670–1688.e7.

32. Sansone R, Sansone L. Doctor Shopping: A Phenomenon of Many Themes. Innovations in clinical neuroscience 2012;9:42–46.

33. Vernon-Roberts A, Safe M, Day AS. Editorial: Pediatric Functional Gastrointestinal Disorders: Challenges in Diagnosis and Treatment. Gastrointestinal Disorders 2024;6:308–312.

34. Daly M, Zarate-Lopez N. Functional gastrointestinal disorders: History taking skills in practice. Clinical Medicine 2021;21:e480–e486.

35. Brekke M, Brodwall A. Understanding parents’ experiences of disease course and influencing factors: a 3-year follow-up qualitative study among parents of children with functional abdominal pain. BMJ Open 2020;10:e037288.

36. Brodwall A, Brekke M. General practitioners’ experiences with children and adolescents with functional gastrointestinal disorders: a qualitative study in Norway. Scand J Prim Health Care 2021;39:543–551.

37. Tome J, Kamboj AK, Loftus CG. Approach to Disorders of Gut-Brain Interaction. Mayo Clinic Proceedings 2023;98:458–467.

38. Hale AE, Smith AM, Christiana JS, et al. Perceptions of Pain Treatment in Pediatric Patients With Functional Gastrointestinal Disorders. The Clinical journal of pain 2020;36:550–557.

39. Burgell RE, Hoey L, Norton K, et al. Treating disorders of brain-gut interaction with multidisciplinary integrated care. Moving towards a new standard of care. JGH Open 2024;8:e13072.

40. Beinvogl B, Burch E, Snyder J, et al. Multidisciplinary Treatment Reduces Pain and Increases Function in Children With Functional Gastrointestinal Disorders. Clinical gastroenterology and hepatology 2019;17:994–996.

41. Wehi P, Roa T. Reciprocal relationships: identity, tradition and food in the Kīngitanga Poukai He Manaakitanga: o te tuakiri, o te tikanga me te kai ki te Poukai o te Kīngitanga. . 10.31235/osf.io/tz746. SocArXiv Preprint 2019.

